# Peripheral and central contributions to persistent pain in rheumatoid arthritis: an unbiased latent profile analysis identifies four mechanism-based phenotypes

**DOI:** 10.64898/2026.06.24.26356419

**Authors:** Zoe Rutter-Locher, L. Zhao, Sam Norton, Leonie S. Taams, Bruce W. Kirkham, Kirsty Bannister

## Abstract

**Background:** Pain frequently persists in rheumatoid arthritis (RA), despite effective control of inflammation. The mechanisms driving this residual pain remain poorly characterised in individual patients.

**Methods:** In 172 patients with established RA and clinically relevant pain (mean NRS 6.5/10) and 80 pain-free controls, we combined indicators of inflammatory disease (CRP, joint counts, power Doppler ultrasound), centrally mediated pain (Widespread Pain Index, painDETECT), psychological distress (PHQ-ADS) and quantitative sensory testing (QST). Latent profile analysis was applied without predefined thresholds.

**Results:** Four phenotypes were identified: a peripheral, low-inflammation/low-central phenotype (38%); a predominantly inflammatory phenotype (7%); and moderate (43%) and severe (12%) centrally mediated phenotypes. Centrally mediated phenotypes reported the highest pain (NRS 8.2), worst disease impact and lowest employment. DAS28-CRP was similar in both the inflammatory and severe centrally mediated phenotypes but for different reasons - swollen joints and CRP versus tender joints - and did not distinguish them. Conditioned pain modulation was impaired relative to controls (p<0.001) and most reduced in the severe centrally mediated phenotype. Psychological distress was the strongest independent predictor of pain severity (model R²=0.33), whereas inflammatory markers were not. Principal components analysis identified swollen joint count (loading 0.63) and the tender–swollen joint difference (loading 0.60) as accessible clinical markers of the inflammatory and centrally mediated phenotypes respectively.

**Conclusions:** A data driven approach identified four mechanism-based pain phenotypes in RA. This framework moves pain assessment beyond inflammation alone and provides a basis for testing analgesic strategies to target the predominant pain mechanism in individual patients.

## 1. Introduction

Pain remains one of the most disabling symptoms for people living with rheumatoid arthritis (RA), despite the widespread use of highly effective disease-modifying therapies[47]. This residual pain likely reflects non-inflammatory mechanisms in a substantial proportion of people[43]. Disease activity in RA is typically monitored using composite scores such as disease activity score in 28 joints using C-Reactive Protein (DAS28-CRP), which incorporate tender and swollen joint counts, CRP and patient global assessment, and guide decisions about escalating immunosuppression. When non-inflammatory pain is not recognised, the resulting high composite disease activity scores can be misattributed to active disease, inflating these scores [16,28] and prompting escalation of immunosuppression that is unlikely to relieve it.

Pain phenotyping, where patients are grouped by biological and clinical pain profiles may allow potential mechanism-targeted analgesic treatment. Features consistent with centrally mediated pain, including widespread pain and heightened pain sensitivity, have been reported in up to 40% of people with RA[37], and approximately 25% have been proposed to experience pain sensitisation driven by peripheral processes in the absence of detectable inflammation[4]. Psychological distress, including depression and anxiety, is highly prevalent in RA[26,48] and shares supraspinal pain-processing circuits with centrally mediated pain[3,8], yet is rarely incorporated into phenotyping frameworks alongside inflammatory and sensory measures. Identifying which mechanism predominates in an individual patient could match treatment to the underlying biology and enable phenotype-stratified clinical care.

Existing phenotyping approaches have important limitations. No single measure captures the constructs of inflammation or centrally mediated pain in full[45], and all infer rather than measure them directly[11,31]. Multiple measures are therefore needed to characterise pain phenotypes more completely, yet most studies rely on a single instrument, typically a questionnaire[37] or sensory testing[46]. Many studies also apply cut-offs that have not been validated in RA, which introduces assumptions about what constitutes a given phenotype in this population. Finally, because few studies incorporate an objective measure of inflammatory burden, the contribution of inflammation cannot reliably be distinguished from that of centrally mediated pain or peripheral sensitisation.

We therefore took a data-driven approach. In a large cohort of patients with established RA and clinically relevant pain (NRS ≥3), we combined objective measures of inflammatory burden such as ultrasound, patient-reported features of centrally mediated pain, psychological distress, and static and dynamic quantitative sensory testing (QST), together providing mechanistic data alongside patient reported measures. Such comprehensive assessment is, however, impractical for routine clinical use, in which long questionnaires and QST require specialist time and equipment. Identifying a small number of clinically accessible markers that are most representative of the resulting phenotypes is therefore an equally important step towards translation. Using latent profile analysis, an unsupervised method that derives subgroups directly from the data without predefined thresholds, we aimed to (i) identify pain phenotypes in RA from inflammatory, sensory and psychological markers; (ii) characterise the patient-reported and sensory profile distinguishing these phenotypes; (iii) determine which features contribute most to pain severity; and (iv) identify clinically accessible screening markers of these phenotypes suitable for routine practice.

## 2. Methods

### 2.1 Design and participants

This was a cross-sectional, observational study of pain mechanisms in RA. Patients were recruited from rheumatology outpatient clinics at Guy’s Hospital and King’s College Hospital, London (REC references 21/LO/0712 and 24/LO/0046). Eligible patients had established RA (time since diagnosis ≥ 1 year) and an average numeric rating scale (NRS) pain score ≥3/10 over the preceding four weeks. Exclusion criteria were age under 18 years, inability to consent or adhere to protocols, pregnancy or breastfeeding, non-RA immunosuppression, recent investigational agents, and severe peripheral vascular disease or peripheral neuropathy. Opioids, gabapentin and pregabalin were withheld for 24 hours before assessment. Pain-free controls (NRS 0/10; KCL reference HR/DP-22/23-34222) were recruited via King’s College London and underwent the same exclusions. All participants gave written informed consent.

### 2.2 Assessments

Inflammatory disease activity was assessed by CRP, 28-joint tender and swollen joint counts (TJC, SJC), DAS28-CRP, and power Doppler musculoskeletal ultrasound (OMERACT-graded, 12 joints). Features of centrally mediated pain and psychological distress were captured by the Widespread Pain Index (WPI), the painDETECT questionnaire and a combined Patient Health Questionnaires Anxiety and Depression Scale (PHQ-ADS score;PHQ-8 for depression + GAD-7 for anxiety). Disease impact was measured with the Rheumatoid Arthritis Impact of Disease (RAID) score.

Static QST followed German Research Network on Neuropathic Pain (DFNS) protocols[36]. Mechanical detection threshold (MDT) and mechanical pain threshold (MPT) were assessed with standardised von Frey filaments and weighted pinprick stimulators, and wind-up ratio (WUR) as the ratio of perceived intensity to a single versus a train of repeated pinprick stimuli. These were measured at an RA-affected joint and the contralateral forearm (non-articular control site). Pressure pain threshold (PPT) was measured by handheld algometry at an affected joint and at the bilateral trapezius (non-articular control site). Values were normalised to age-, sex- and site-matched control reference data by z-transformation: (participant value − control mean) / control SD. A z-score ≥ +1.96 SD defined a participant as “sensitive” at that measure and site.

Dynamic QST used computer-controlled cuff algometry on the lower leg[13]. Pain detection threshold (PDT) and pain tolerance threshold (PTT) were recorded as the pressures eliciting first pain and maximally tolerated pain. Temporal summation of pain (TSP) was assessed during repeated cuff stimulation, with pain rated on a 0–10 visual analogue scale (VAS). TSP was quantified as the absolute change between the mean VAS of stimuli 8–10 and stimuli 1–3. Participants were classified as TSP facilitators if their TSP exceeded the upper bound of the 95% confidence interval of the control distribution (3.1)[34]. Conditioned pain modulation (CPM) was assessed using a contralateral conditioning cuff stimulus, with the CPM effect quantified as the change in PDT (and PTT) during versus before conditioning. Full protocols have been published previously[39]. The CPM effect was defined as PDT (or PTT) during conditioning minus baseline. CPM non-responders were defined two ways: Definition 1, a CPM effect ≤20% of the baseline value[13]; and Definition 2, a CPM effect below the lower bound of the 95% confidence interval of the control distribution (8 for PDT, 8.8 for PTT)[34].

Controls were assessed for age, sex and ethnicity and underwent the same PHQ-8, GAD-7 and QST protocols at the same sites.

### 2.3 Latent profile analysis

Latent profile analysis identified subgroups based on differing patterns of phenotypic markers of inflammation (CRP, SJC, power Doppler score), centrally mediated pain (WPI, painDETECT, trapezius PPT) and psychological distress (PHQ-ADS). Skewed inflammatory variables were transformed (CRP: log-transformed; joint counts and ultrasound variable: square-root transformed) and all indicators z-standardised. Models of 1–6 profiles were estimated by generalised structural equation modelling, using the STATA gsem command, and compared using the Bayesian and Akaike information criteria, profile separation, minimum profile size and clinical interpretability (Table S1). Each participant was assigned to the profile with the highest posterior probability.

Group-level QST in RA versus controls was compared by t-tests. Standardised (z-score) static QST values were derived relative to age-, sex- and site-matched control reference data, with z ≥ +1.96 defining “sensitive.” TSP facilitators and CPM non-responders were defined relative to the control distribution (Supplementary Methods). Differences across latent phenotypes in patient-reported and QST measures were tested by linear regression adjusted for age and sex, with Bonferroni-corrected pairwise comparisons of the adjusted means; categorical variables were compared by χ². Independent predictors of pain severity (NRS average pain) were examined by multivariable linear regression on standardised candidate measures (WPI, painDETECT, PHQ-ADS, trapezius PPT, CRP, power Doppler, TJC, SJC adjusted for age, sex). Analyses used Stata 17.0 and R; p < 0.05 was considered significant.

To identify clinically accessible markers of the phenotypes, principal components analysis (PCA) was applied separately to inflammatory indicators (CRP, power Doppler, SJC, TJC) and to centrally mediated indicators (WPI, painDETECT, PHQ-ADS, trapezius PPT); the tender–swollen joint difference (T–SJC), routinely recorded in clinical practice, was then added to the centrally mediated model. The first principal component was retained with component loadings and uniqueness used to identify the measures most representative of the underlying construct, interpreting loadings ≥0.55 as good and ≥0.63 as very good.

### 2.4 Sample size

This study is an exploratory secondary analysis with no formal sample size calculation conducted for the latent profile analysis. The initial sample size target of 60–100 patients per recruiting site was based on a two-group comparison of QST measures between RA patients and pain-free controls (target >60). This would allow subgroup analysis by patients with versus without clinically relevant inflammation. Based on the rule of thumb that stable profile recovery requires approximately 30–50 participants per profile[33], a sample of 172 patients would support identification of four to five profiles if profiles were roughly equal in size. Profiles with fewer than 30 participants are interpreted with caution. As this was an exploratory study, the significance threshold was fixed at 5% without correction across the multiple outcomes tested; Bonferroni correction was applied only to the post-hoc pairwise comparisons between phenotypes within each outcome.

## 3. Results

### 3.1 Cohort and group-level sensory profile

Of 252 participants, 172 had RA (mean age 54 years, 85% female, 59% White British) and 80 were pain-free controls. Mean DAS28-CRP was 4.1 and mean NRS pain 6.5/10; power Doppler activity was present in at least one joint in 56%. Patient-reported features of centrally mediated pain and psychological distress were common: 49% met widespread-pain criteria (WPI ≥7), 41% had neuropathic-like symptoms on painDETECT, and 60% and 75% screened positive for anxiety and depression respectively, with both significantly higher than in controls (Table 1). At the group level, RA patients showed widespread cutaneous (MPT) and deep-tissue (PPT) sensitisation (Z score ≥1.96 at joint and non-articular site). CPM effect was reduced relative to controls (p < 0.001), whereas mean TSP did not differ (p = 0.28), although 40% of patients met the control-referenced threshold for TSP facilitation (Table 1). Full demographics and QST values are shown in Supplementary material Table S1.

**Table 1.** Cohort characteristics and group-level quantitative sensory testing. Values are mean (SD) or n (%). †Controls were significantly younger and included fewer women; full demographics in Table S3. “Sensitive” denotes a z-score ≥ +1.96 versus matched controls. **Abbreviations:** DAS28, 28-joint Disease Activity Score; NRS, numeric rating scale; MPT, mechanical pain threshold; PPT, pressure pain threshold; PDT, pain detection threshold; PTT, pain tolerance threshold; TSP, temporal summation of pain; CPM, conditioned pain modulation.

| Characteristic | RA (n=172) | Controls (n=80) | P |
| --- | --- | --- | --- |
| **Demographic and disease characteristics** |  |  |  |
| Age, years, mean (SD) | 54 (14) | 41 (15) | <0.05 |
| Female, n (%) | 146 (85) | 51 (64) | <0.05 |
| DAS28-CRP, mean (SD) | 4.1 (1.0) | — | — |
| Tender joint count (28), mean (SD) | 9.5 (7) | — | — |
| Swollen joint count (28), mean (SD) | 2.2 (3) | — | — |
| Power Doppler $\geq 1$ joint, n (%) | 96 (56) | — | — |
| NRS average pain (0–10), mean (SD) | 6.5 (2) | 0 (0) | — |
| **Patient-reported pain and distress** |  |  |  |
| Widespread Pain Index $\geq 7$, n (%) | 84 (49) | — | — |
| painDETECT neuropathic-like (Score $\geq 19$), n (%) | 70 (41) | — | — |
| GAD-7 $\geq 5$ (anxiety), n (%) | 103 (60) | 27 (34) | <0.001 |
| PHQ-8 $\geq 5$ (depression), n (%) | 129 (75) | 22 (28) | <0.001 |
| **Group-level quantitative sensory testing** |  |  |  |
| MPT, forearm / joint, mean (SD) | 2.0 (3), 2.4 (3) | — | — |
| PPT, trapezius / joint, mean (SD) | 1.5 (1), 2.4 (2) | — | — |
| Cuff PDT, kPa, mean (SD) | 17.8 (9.0) | 19.2 (9.0) | 0.26 |
| Cuff PTT, kPa, mean (SD) | 34.0 (13.7) | 45.0 (20) | <0.001 |
| TSP (difference), mean (SD) | 2.0 (1.9) | 2.4 (3.0) | 0.28 |
| TSP facilitators, n (%) | 61/154 (40) | 16/80 (20) | — |
| CPM effect, PDT, mean (SD) | 4.8 (8) | 10.3 (10.4) | <0.001 |
| CPM effect, PTT, mean (SD) | 4.6 (7) | 10.8 (9.0) | <0.001 |

### 3.2 Four data-driven pain phenotypes

Latent profile analysis supported a four-profile solution as the best balance of fit and clinical interpretability (lowest BIC; Tables S2 and S3). The profiles were defined by their standardised indicator means (Figure 1, Table S4). Profile 1 (38%) showed low or near-average values across inflammatory, centrally mediated and psychological indicators. Profile 2 (7%) was defined by high inflammatory activity (CRP, power Doppler, SJC) with low centrally mediated and psychological features. Profile 3 (43%) showed moderate elevations in centrally mediated pain and distress with low inflammation, and Profile 4 (12%) showed very high centrally mediated pain features, distress, pressure-pain sensitisation and tender joint counts, with low inflammation apart from modestly raised CRP. We interpret these as a peripheral non-inflammatory phenotype, a predominantly inflammatory phenotype, and moderate and severe centrally mediated phenotypes, respectively.

**Figure 1.**
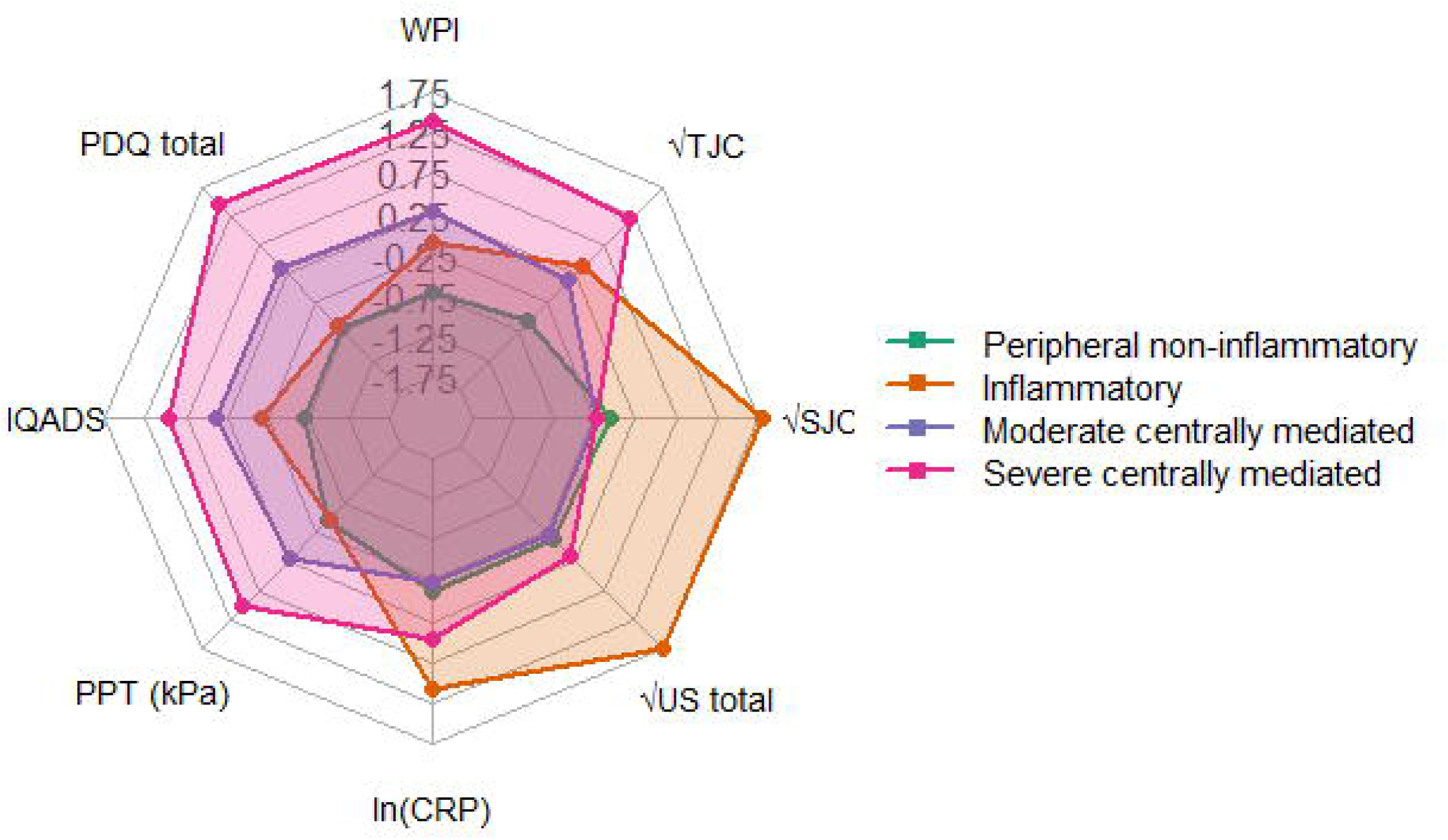
Latent profile analysis of inflammatory, centrally mediated and psychological indicators. Radar plot of standardised (z-score) indicator means across the four phenotypes; coloured polygons represent mean profile values, with each axis a z-score (trapezius PPT reversed to align directionality).

### 3.3 Pain, disease impact and sensory profile by phenotype

There was no difference in age, sex, ethnicity or disease duration across phenotypes (Table 2). Pain severity and disease impact increased stepwise from profile 1 to 4, and were worst in the severe centrally mediated phenotype, which reported the highest pain (NRS 8.2), the worst scores across all RAID domains (Table S5) and the lowest employment. The peripheral phenotype had the lowest burden, yet mean pain (5.4) and RAID (4.5) remained above acceptable thresholds. DAS28-CRP was similarly high in the inflammatory and severe centrally mediated phenotypes, driven by different components in each; high swollen joint count and CRP in the inflammatory phenotype, but high tender joint count with low swollen joint count in the severe centrally mediated phenotype.

**Table 2.**
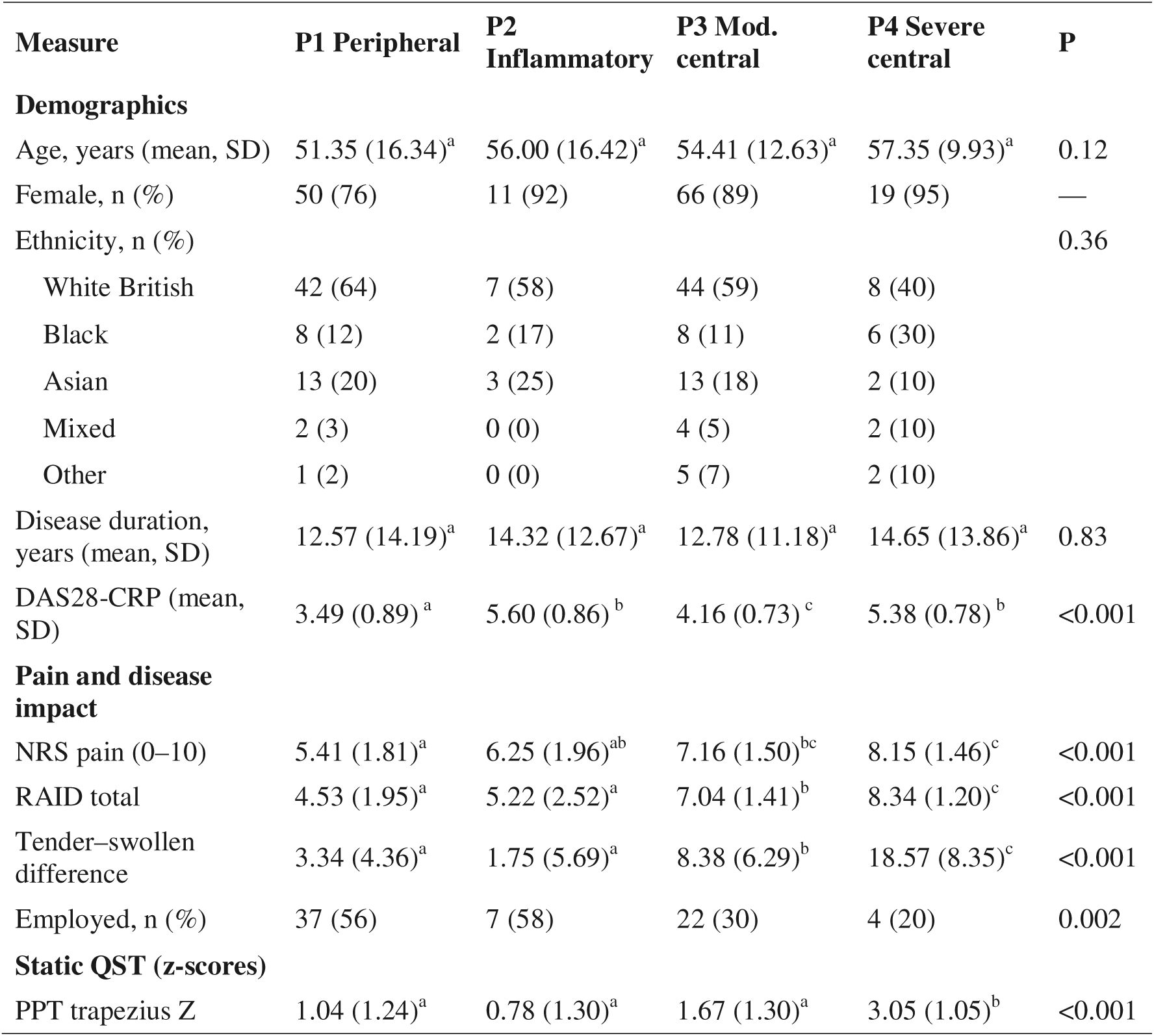
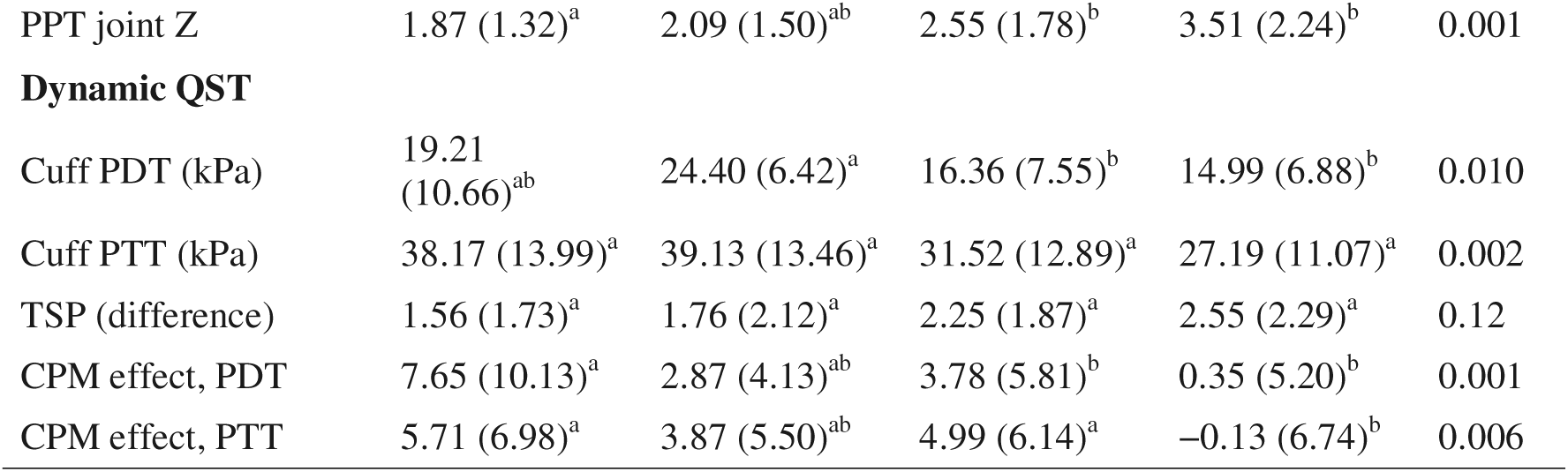
Demographics, pain, disease impact and sensory profile by latent phenotype. Values are mean (SD) unless stated; sex, ethnicity and employment are n (%). P from regression (continuous outcomes) or chi-square (categorical), adjusted for age and sex where applicable. Superscripts (a, b, c) denote Bonferroni-adjusted pairwise differences; groups sharing a letter do not differ significantly. RAID domain scores and full static QST are in Table S5 and S6. RAID, Rheumatoid Arthritis Impact of Disease; QST, quantitative sensory testing; PPT, pressure pain threshold; PDT, pain detection threshold; PTT, pain tolerance threshold; TSP, temporal summation of pain; CPM, conditioned pain modulation.

| Measure | P1 Peripheral | P2 Inflammatory | P3 Mod. central | P4 Severe central | P |
| --- | --- | --- | --- | --- | --- |
| **Demographics** |  |  |  |  |  |
| Age, years (mean, SD) | 51.35 (16.34)<sup>a</sup> | 56.00 (16.42)<sup>a</sup> | 54.41 (12.63)<sup>a</sup> | 57.35 (9.93)<sup>a</sup> | 0.12 |
| Female, n (%) | 50 (76) | 11 (92) | 66 (89) | 19 (95) | — |
| Ethnicity, n (%) |  |  |  |  | 0.36 |
| White British | 42 (64) | 7 (58) | 44 (59) | 8 (40) |  |
| Black | 8 (12) | 2 (17) | 8 (11) | 6 (30) |  |
| Asian | 13 (20) | 3 (25) | 13 (18) | 2 (10) |  |
| Mixed | 2 (3) | 0 (0) | 4 (5) | 2 (10) |  |
| Other | 1 (2) | 0 (0) | 5 (7) | 2 (10) |  |
| Disease duration, years (mean, SD) | 12.57 (14.19)<sup>a</sup> | 14.32 (12.67)<sup>a</sup> | 12.78 (11.18)<sup>a</sup> | 14.65 (13.86)<sup>a</sup> | 0.83 |
| DAS28-CRP (mean, SD) | 3.49 (0.89)<sup>a</sup> | 5.60 (0.86)<sup>b</sup> | 4.16 (0.73)<sup>c</sup> | 5.38 (0.78)<sup>b</sup> | <0.001 |
| **Pain and disease impact** |  |  |  |  |  |
| NRS pain (0–10) | 5.41 (1.81)<sup>a</sup> | 6.25 (1.96)<sup>ab</sup> | 7.16 (1.50)<sup>bc</sup> | 8.15 (1.46)<sup>c</sup> | <0.001 |
| RAID total | 4.53 (1.95)<sup>a</sup> | 5.22 (2.52)<sup>a</sup> | 7.04 (1.41)<sup>b</sup> | 8.34 (1.20)<sup>c</sup> | <0.001 |
| Tender–swollen difference | 3.34 (4.36)<sup>a</sup> | 1.75 (5.69)<sup>a</sup> | 8.38 (6.29)<sup>b</sup> | 18.57 (8.35)<sup>c</sup> | <0.001 |
| Employed, n (%) | 37 (56) | 7 (58) | 22 (30) | 4 (20) | 0.002 |
| **Static QST (z-scores)** |  |  |  |  |  |
| PPT trapezius Z | 1.04 (1.24)<sup>a</sup> | 0.78 (1.30)<sup>a</sup> | 1.67 (1.30)<sup>a</sup> | 3.05 (1.05)<sup>b</sup> | <0.001 |

| **Measure** | **P1 Peripheral** | **P2 Inflammatory** | **P3 Mod. central** | **P4 Severe central** | **P** |
| --- | --- | --- | --- | --- | --- |
| PPT joint Z | 1.87 (1.32)<sup>a</sup> | 2.09 (1.50)<sup>ab</sup> | 2.55 (1.78)<sup>b</sup> | 3.51 (2.24)<sup>b</sup> | 0.001 |
| **Dynamic QST** |  |  |  |  |  |
| Cuff PDT (kPa) | 19.21 (10.66)<sup>ab</sup> | 24.40 (6.42)<sup>a</sup> | 16.36 (7.55)<sup>b</sup> | 14.99 (6.88)<sup>b</sup> | 0.010 |
| Cuff PTT (kPa) | 38.17 (13.99)<sup>a</sup> | 39.13 (13.46)<sup>a</sup> | 31.52 (12.89)<sup>a</sup> | 27.19 (11.07)<sup>a</sup> | 0.002 |
| TSP (difference) | 1.56 (1.73)<sup>a</sup> | 1.76 (2.12)<sup>a</sup> | 2.25 (1.87)<sup>a</sup> | 2.55 (2.29)<sup>a</sup> | 0.12 |
| CPM effect, PDT | 7.65 (10.13)<sup>a</sup> | 2.87 (4.13)<sup>ab</sup> | 3.78 (5.81)<sup>b</sup> | 0.35 (5.20)<sup>b</sup> | 0.001 |
| CPM effect, PTT | 5.71 (6.98)<sup>a</sup> | 3.87 (5.50)<sup>ab</sup> | 4.99 (6.14)<sup>a</sup> | -0.13 (6.74)<sup>b</sup> | 0.006 |

Sensory testing provided mechanistic support for the phenotypes. Cutaneous mechanical hyperalgesia (MPT) was widespread and did not differ between profiles (Table S6). Joint and trapezius pressure-pain sensitivity increased stepwise and was greatest in the severe centrally mediated phenotype, and cuff pain detection and tolerance thresholds were lowest in the two centrally mediated phenotypes, indicating widespread deep-tissue sensitisation. Among dynamic measures, TSP rose numerically across profiles but did not differ significantly (Table 2), including after sensitivity analyses excluding low baseline ratings (Table S6). However, CPM effect reduced across profiles and was lowest, effectively absent or facilitatory, in the severe centrally mediated phenotype, reaching significance for pain-tolerance CPM, indicating impaired descending inhibition as a candidate mechanism in this group (Table 2).

### 3.4 Predictors of pain severity

A multivariable model of standardised measures explained 33% of the variance in pain severity (R² = 0.33, adjusted R² = 0.29; F(10,151) = 7.42, p < 0.001). After adjustment for age and sex, psychological distress was the strongest independent predictor (PHQ-ADS b = 0.042, p = 0.001), followed by neuropathic-like pain (painDETECT b = 0.044, p = 0.048). Inflammatory markers (CRP, power Doppler, SJC), trapezius PPT and WPI were not independently associated with pain severity (all p > 0.1).

### 3.5 Clinical indicators of pain phenotypes

Because the full assessment is impractical in routine clinic, PCA was used to identify accessible markers of each phenotype (Figure 2). For the inflammatory phenotype, a single component explained 42% of the variance, with swollen joint count showing the strongest loading and lowest uniqueness (0.63); given this and its routine availability, SJC was the measure most representative of inflammatory pain, performing comparably to power Doppler ultrasound. For the centrally mediated phenotype, a single component explained 54% of the variance, with painDETECT loading most strongly (0.72). When the routinely recorded tender–swollen joint difference was added, a single component still dominated (50.8% of the variance), painDETECT retained a strong loading (0.70), but T–SJC also loaded at 0.60 indicating that T–SJC, as a routinely available measure, may serve as an accessible clinical indicator of centrally mediated pain.

**Figure 2.**
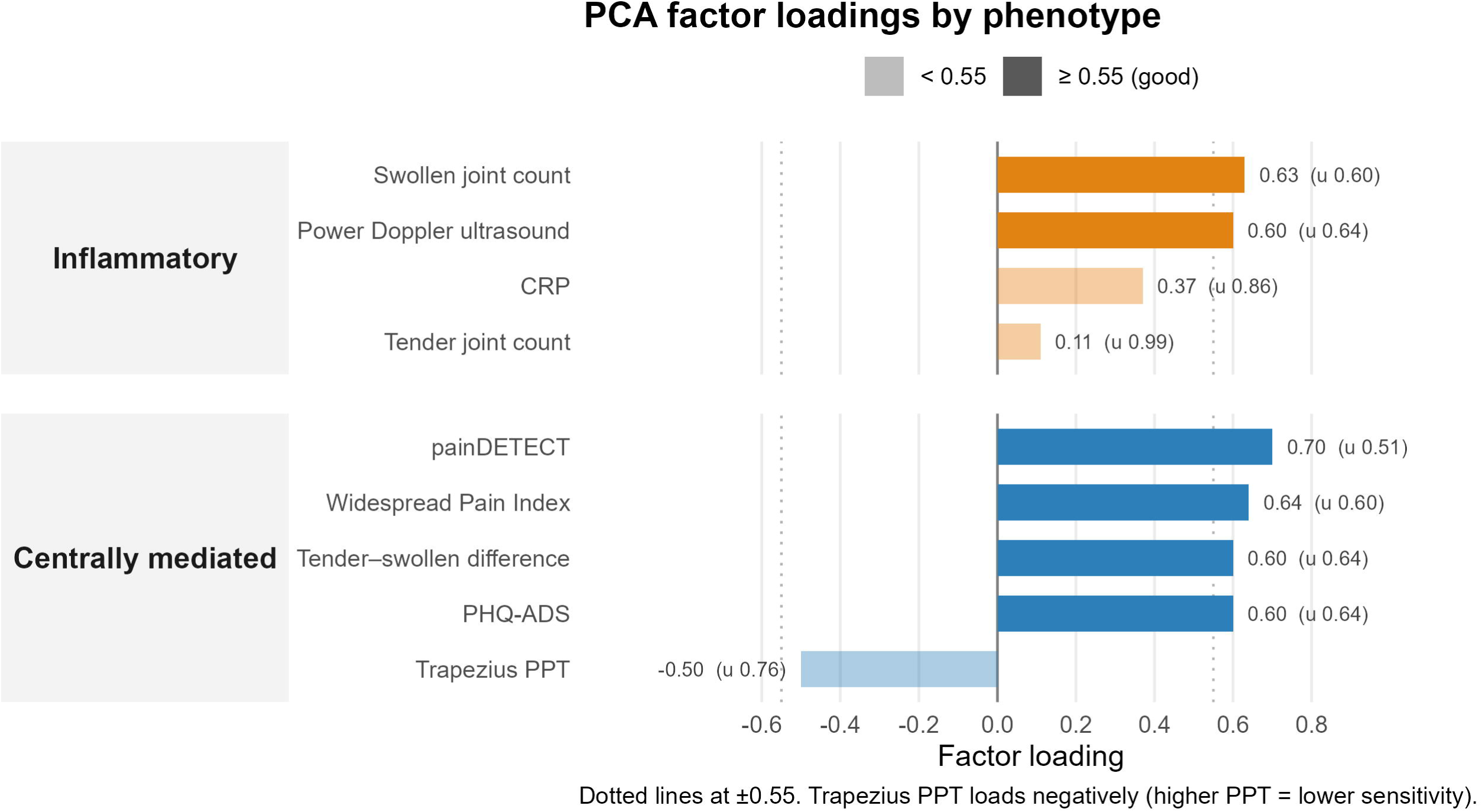
Factor loadings and uniqueness from principal components analysis of inflammatory and centrally mediated indicators. Higher loadings (and lower uniqueness) denote measures more representative of the underlying construct. Trapezius PPT loads negatively because higher pressure-pain thresholds indicate lower sensitivity. T–SJC, tender–swollen joint difference; other abbreviations as Table 1.

## 4. Discussion

By applying latent profile analysis to inflammatory, sensory and psychological markers without predefined thresholds, in consecutive eligible patients and controls, we identified four pain phenotypes directly from the data: a predominantly inflammatory phenotype; moderate and severe centrally mediated phenotypes; and a low-inflammation, low-central-features phenotype with clinically meaningful pain. As in other studies[27,37], the greatest pain and impact of disease burden was carried by the centrally mediated groups, which together comprised over half the cohort. Quantitative sensory testing provided mechanistic support for the phenotypes, and accessible clinical measures, swollen joint count for the inflammatory phenotype and the tender–swollen joint difference for the centrally mediated phenotype, offer a means of identifying them in routine practice. Together, these findings provide an data driven framework for understanding the relative contributions of peripheral and central mechanisms to persistent pain in RA.

The predominantly inflammatory phenotype was defined by high CRP, power Doppler activity and swollen joint counts with minimal centrally mediated features, and was supported by QST showing local joint hypersensitivity without widespread sensitisation, TSP facilitation or impaired CPM. This is consistent with sensitisation of primary afferents innervating the inflamed joint, by pro-inflammatory mediators, including prostaglandins, bradykinin, TNF, IL-6 and nerve growth factor[15,38]. The inflammatory phenotype accounted for only a small proportion of patients and was associated with only moderate pain, reinforcing that in patients with high levels of pain (NRS ≥3 in this study) suppressing inflammation alone is often insufficient to explain, or to relieve, the pain experienced.

The centrally mediated phenotypes carried the greatest burden in terms of pain and impact of disease. For example, sleep disturbance was greatest in the centrally mediated phenotypes, in keeping with literature linking it closely to central pain[20], and may itself be a mediator and a potentially modifiable treatment target[2]. The severe central group also had similarly elevated DAS28 CRP to the inflammatory phenotype, which has direct clinical consequences: patients with centrally mediated pain may be mislabelled as difficult to treat RA[32] or receive escalation of immunosuppression that is unlikely to relieve their pain [18].

The prevalence of centrally mediated features is consistent with previous reports of pain sensitisation [37] and widespread pain in approximately one-third[1,10,28] of patients with RA. Our data extend this literature by incorporating these features within a framework that simultaneously captures inflammatory burden, and by deriving the phenotypes without using thresholds that remain unvalidated in RA. The continuous increase in severity between the moderate and severe centrally mediated phenotypes, and the modestly raised CRP in the severe group, indicates that these phenotypes are not discrete categories but reflect the relative dominance of peripheral and central features existing on a continuum[43].

The dynamic QST paradigms provide mechanistic support for the phenotypes. TSP, a proxy measure of spinal facilitatory processes in which higher values indicate abnormal facilitation[34], was highest in the moderate and severe centrally mediated phenotypes. This increase in TSP did not reach significance, which may reflect the study being underpowered to detect small between-group differences. Floor effect from low baseline stimulus ratings (20% rated the first stimulus 0/10) may also contribute, although sensitivity analyses excluding these participants produced similar results. CPM was impaired in RA overall relative to controls, in keeping with another large controlled study [20], and was most reduced in the severe centrally mediated phenotype, reaching significance for pain tolerance. This suggests impaired descending inhibitory control, mediated by brainstem noradrenergic and serotonergic projections, as a mechanism to target with agents such as serotonin–noradrenaline reuptake inhibitors. The convergence of impaired descending inhibition with high psychological distress in the severe centrally mediated phenotype is consistent with shared supraspinal circuitry: the anterior cingulate cortex, insula and periaqueductal grey modulate both nociceptive transmission and affective processing, and noradrenergic and serotonergic projections from the brainstem are disrupted in states of chronic pain and depression alike[3]. It is important to note that conclusions regarding CPM and TSP apply at the group level. Both show substantial inter-individual variability[34] and only modest test–retest reliability[6,44] so in their current form they cannot reliably classify individual patients for treatment selection, or predict an individual’s pain trajectory or response to treatment.

Consistent with this overlap, psychological distress was the single strongest independent predictor of pain severity and was closely associated with other centrally mediated pain features across phenotypes. Given the growing evidence for a bidirectional relationship between psychological distress and pain[9,41], this supports screening for, and targeted management of, depression and anxiety, and identifies descending inhibitory pathways as a candidate target for pharmacological intervention, for example with serotonin–noradrenaline reuptake inhibitors.

The last phenotype reported pain and RAID scores which were above acceptable thresholds[30] but were not explained by either detectable inflammation or centrally mediated features. We therefore interpret this phenotype as one in which pain is driven by other peripheral processes, which could include clinically and ultrasonographically undetectable inflammation (persistence of tissue-resident cells and mediators at subclinical levels, for example synovial fibroblasts producing IL-6 and leukaemia inhibitory factor[5,21,22,38]), and/or peripheral sensitisation (such as lasting changes in nociceptor excitability after prior inflammation, including altered sodium-channel and TRPV1 function[38,40] and the awakening of silent nociceptors). We previously proposed a peripherally mediated pain phenotype in RA[4]; in this analysis we identify a similar group in a population assessed for pain mechanisms in a data-driven manner, without the predefined cut-offs used previously. This phenotype cannot be confirmed by any single test and must be inferred; distinguishing it definitively from a simply lower-burden group would require other approaches such as microneurography[25] and exploration of underlying driving mediators in the synovial fluid and serum. This population will be important to identify; these patients are unlikely to respond to either disease-modifying drugs or from centrally acting analgesics, but identification could aid the translational discovery of peripheral analgesic targets.

Identifying these phenotypes in clinic requires an accessible, practical tool. In our analysis, swollen joint count best represented the inflammatory phenotype, and the tender–swollen joint difference best represented the centrally mediated phenotype. The tender–swollen difference is particularly attractive: it is already recorded routinely, has been proposed as a marker of fibromyalgic RA[35], is associated with lower ultrasound-detected synovitis[29] and has recently been shown to be associated with QST measures[7,17,23]. It has not, however, been developed as a measure of centrally mediated pain; tenderness reflects several processes, swollen joint counts are observer-dependent, and no diagnostic threshold has been established. It is therefore best regarded as a continuous screening measure that flags patients for formal assessment of centrally mediated pain, rather than as a stand-alone diagnostic test.

A strength of this study is its data-driven, multi-measure design. Previous data-driven phenotyping in RA has used clinical and self-report variables alone, such as hierarchical clustering of symptom reports [19] and latent class analysis of disease-activity components[27]. Using multiple complementary measures mitigates the limitations of any single proxy; no single questionnaire or QST measure captures centrally mediated pain completely[45], and by not using predefined cut-offs we avoid applying thresholds not validated in this population.

Several limitations should be considered. All indicators are proxies; there is no direct way of measuring of inflammation or centrally mediated pain available. Latent profile solutions depend on the variables included and on sample size; although 172 patients supported a clinically coherent four-profile solution, the small inflammatory profile (n=12) falls below the thresholds for stable estimation and therefore requires replication. Furthermore, smaller subgroups may have gone undetected and the latent profile analysis solution should be regarded as hypothesis generating. Controls were younger and included fewer men, which may exaggerate group-level QST differences[12–14,24], although age and sex were adjusted for in all phenotype comparisons. Finally, separating overlapping mechanisms is inherently artificial, and contributors such as sleep and social context were not fully modelled[20,42,43]. External validation in larger, independent cohorts is required.

In conclusion, an unbiased, data-driven approach identified four pain phenotypes in RA, supported by mechanistic evidence. This framework moves pain assessment in RA beyond inflammation alone and provides a foundation for two translational goals: testing in future trials whether analgesic strategies can be matched to the predominant pain mechanism and identifying the cellular and molecular mediators, particularly of peripheral sensitisation, that could support studies to identify potential novel analgesic targets.

## Declarations

### Data availability

The data and the analysis code underlying this article will be shared on reasonable request to the corresponding author.

### Patient and public involvement

Patients and the public were involved throughout this research, from shaping the research question at a workshop with expert patients, clinicians, and academics, where pain was identified as the top priority in inflammatory arthritis, to confirming its importance at a subsequent public engagement event. Patients contributed to study design, with National Rheumatoid Arthritis Society members reviewing pain questionnaires and two patient research partners (PRPs) refining the protocol, consent forms, and information materials. PRPs were also involved in interpreting findings, ensuring conclusions reflected patient perspectives, and providing feedback on manuscript wording. Results have been shared with patients and the public through lay summaries and presentations at patient engagement events.

